# Medication Management of Anxiety and Depression by Primary Care Pediatrics Providers: A Retrospective Electronic Health Record Study

**DOI:** 10.1101/2021.04.11.21255276

**Authors:** Talia R. Lester, Yair Bannett, Rebecca M. Gardner, Heidi M. Feldman, Lynne C. Huffman, Stanford University School of Medicine

## Abstract

**Objectives:** To describe medication management of children diagnosed with anxiety and depression by primary care providers.

**Study Design/Methods:** We performed a retrospective cross-sectional analysis of electronic health record (EHR) structured data. All visits for pediatric patients seen at least twice during a four-year period within a network of primary care clinics in Northern California were included. Descriptive statistics summarized patient variables and most commonly prescribed medications. For each subcohort (anxiety, depression, and both (anxiety+depression)), logistic regression models examined the variables associated with medication prescription.

**Results:** Of all patients (N=93,025), 2.8% (n=2635) had a diagnosis of anxiety only, 1.5% (n=1433) depression only, and 0.79% (n=737) both anxiety and depression (anxiety+depression); 18% of children with anxiety and/or depression had comorbid ADHD. A total of 14.0% with anxiety (n=370), 20.3% with depression (n=291), and 47.5% with anxiety+depression (n=350) received a psychoactive non-stimulant medication. For anxiety only and depression only, sertraline, citalopram, and fluoxetine were most commonly prescribed. For anxiety+depression, citalopram, sertraline, and escitalopram were most commonly prescribed. The top prescribed medications also included benzodiazepines. Logistic regression models showed that older age and having developmental or mental health comorbidities were independently associated with increased likelihood of medication prescription for children with anxiety, depression, and anxiety+depression. Insurance type and sex were not associated with medication prescription.

**Conclusions:** PCPs prescribe medications more frequently for patients with anxiety+depression than for patients with either diagnosis alone. Medication choices generally align with current recommendations. Future research should focus on the use of benzodiazepines due to safety concerns in children.

## Background

According to recent estimates, one in six children aged 6 to 17 have a treatable mental health disorder.^1^ Approximately 75% of adult mental health disorders have onset in childhood. ^2,3^ Thus, the burden of mental health disorders in the pediatric population has long lasting implications. In adulthood, mental health conditions can lead to decreased work productivity, relationship problems, and overall decreased quality of life. Mental health conditions also represent a significant financial burden on the US economy.^4,5^ Evidence suggests that early treatment of mental health disorders leads to better outcomes.^6,7^ Despite the evidence that early detection and treatment of pediatric mental health conditions is beneficial for both individuals and society, studies suggest that many children with mental health conditions are not receiving adequate care.^8^ Barriers to care are numerous and include inadequate primary care provider training and limited access to subspecialists.^9,10^

Depression and anxiety occur in 3 and 7% of pediatric patients, respectively, in the United States.^11^, and most children with these conditions present first to a primary care provider (PCP).^2^ Standard of care for childhood anxiety and depression is a multimodal approach, including pharmacological and non-pharmacological treatments. Current guidelines indicate that medication should be considered in moderate and severe cases.^12–14^ When medication is indicated for pediatric patients with anxiety or depression, the selective serotonin reuptake inhibitor (SSRI) class of medication has a substantial evidence base.^12,15,16^ Among the SSRIs, fluoxetine has the most evidence for good response for both anxiety and depression.^15^ For anxiety, sertraline and fluvoxamine also have high-quality multisite studies demonstrating efficacy and safety in children.^15,16^ For major depressive disorder, fluoxetine is FDA-approved for children, and fluoxetine and escitalopram both are approved for adolescents.

Although most children with anxiety and depression present first to their primary care provider (PCP), the extant literature lacks objective data on the clinical care provided for children with anxiety and depression by PCPs. Prior studies investigating the role of primary care providers in managing patients with anxiety and depression have utilized provider report to assess comfort level and prescribing practices. Stein et al surveyed pediatricians and found that 80% agreed that pediatricians should be able to identify conditions of attention-deficit/hyperactivity disorder, eating disorders, substance use, and behavior problems. 88% agreed that pediatricians should be identifying depression, but only 25% felt pediatricians should be responsible for treating and managing depression.^17^ Response percentages were similar for pediatric anxiety. To date, there is a gap in the extant literature that reflects limited understanding of what primary care providers do in practice (i.e., provider behavior) when caring for children with anxiety and depression, including rates of prescribing medication and types of medications used. Pediatric patients may be prescribed psychoactive medications that lack evidence for use in the pediatric population. For example, Bushnell et al found that 38% of children with benzodiazepine prescription did not have an appropriate indication.^18^

In this study, we used EHR data to describe medication management for anxiety and depression by pediatric PCPs. We also identified patient characteristics associated with prescription of medication. We hypothesized that older age and higher number of comorbidities would be associated with likelihood to be prescribed medication for anxiety or depression.

## Methods

### Setting and Population

Packard Children’s Healthcare Alliance (PCHA) is a community-based network of 25 primary care offices in Northern California, affiliated with Lucille Packard Children’s Hospital and Stanford Children’s Health. The Stanford University School of Medicine institutional review board (IRB) determined that this study was not human subject research.

### Data Sources

We conducted a retrospective review of electronic health records for a cohort including all pediatric patients seen at PCHA clinics for at least two visits during a four-year period (October 1, 2015 to September 30, 2019). Deidentified structured data from all office encounters were included. Supplement 1 shows the study cohort flow-chart. Initial review included patients aged 0-21 years. The subsequent logistic regression analysis included only patients 6-18 years, as only a few patients with anxiety and/or depression diagnoses outside this age range received medication. Patient data included age in years, sex, race, ethnicity, and medical insurance.

### Measures

#### Identifying patients with anxiety and depression

Patients with anxiety and/or depression were identified based on the presence of at least one ICD-10 visit diagnosis code associated with anxiety or depression. Both disorder-level codes (“F codes”, for example: “Generalized Anxiety Disorder”) and symptom-level codes (“R codes”, for example: “worries”) were included. See Supplement 2 for a full list of ICD-10 codes used. Once selected for inclusion, patients were defined as an “anxiety patient” if they had only an anxiety diagnosis (with no depression), “depression patient” if they had only a depression diagnosis (with no anxiety) or “anxiety+depression patient” if they had both anxiety and depression diagnoses during the study period, although the two diagnoses were not required to be present at the same visit.

#### Defining Medication List

A list of all medications prescribed during the study period at visits with anxiety and/or depression diagnoses was reviewed, and a comprehensive list of psychoactive medications (excluding stimulants) was generated.

#### Defining Comorbidities

Behavioral and mental health comorbidities were identified based upon the presence of ICD-10 diagnosis codes associated with the following: Attention-deficit/hyperactivity disorder (ADHD), Sleep Disorders, Trauma and Stressor Related Disorders, Autism Spectrum Disorders (ASD), Obsessive Compulsive and Related Disorders, Feeding and Eating Disorders, Disruptive, Impulse Control, and Conduct Disorders, Substance-Related and Addictive Disorders, and Bipolar and Related Disorders. These categories were chosen based upon broad categories in the Diagnostic and Statistical Manual of Mental Disorders, 5^th^ Edition (DSM-V).^19^ The comorbid diagnosis was not required to be present at the same visit as the anxiety and/or depression diagnosis.

### Statistical Analyses

#### Descriptive statistics

For patients with anxiety and/or depression, frequency counts, proportions, means and standard deviations were used to summarize data representing patient characteristics (demographics, comorbidities) and most commonly prescribed medications.

#### Logistic regression models

Three logistic regression models addressing 1) anxiety, 2) depression and 3) anxiety+depression were performed to determine association of patient characteristics with likelihood to receive medication. Predictor variables included: age group (6-12 and 13-18), sex, insurance (private, public, military, or unknown), number of mental health and/or behavioral comorbidities (0, 1, or 2+), specific comorbid diagnosis (ADHD, ASD, sleep disorder, or trauma and stressor related disorders), and referral to Developmental-Behavioral Pediatrics or Psychiatry services within Stanford Children’s Health. These specific comorbid conditions were selected because they were the most common among patients in the data set. Patients under 6 years of age were excluded from the models due to infrequent medication use in this age group (Supplement 3). We calculated adjusted odds ratios (aOR) and 95% confidence intervals (CI) from the regression models.

Since there was a high rate of missing race and ethnicity data in our cohort (approximately 30%), race and ethnicity were not included in the regression models. Imputation was not attempted since missing data was highly dependent on practice/clinic. To explore the potential effect of race and/or ethnicity on medication use for anxiety and depression, we determined the prevalence of psychoactive medication use in patients with anxiety and/or depression in each race and ethnicity category only for cases where the race/ethnicity data were complete (non-missing). We used chi-square tests to compare proportions.

Data cleaning and reformatting were performed using R version 3.6.2. All analyses were conducted using SPSS 26 and 27.^20^ All statistical tests were two-sided and conducted at the 0.05 significance level.

## Results

### Patient Characteristics and Prevalence of Anxiety and/or Depression

Of 93,025 patients 0-21 years old seen at least twice during the study period for any reason, 4805 (5.2%) received a diagnosis of anxiety and/or depression.

Of the 4805, 41.7% were male, 40.1% were white, and 76.0% were privately insured. Table 1 describes characteristics of the 4805 patients: 2635 (54.8%) had at least one visit with a diagnosis of anxiety only (symptom or disorder level), n=1433 (29.8%) had at least one visit with a diagnosis of depression only (symptom or disorder), and n=737 (15.3%) had a diagnoses of both anxiety and depression (anxiety+depression) (symptom or disorder) during the study period.

**Table 1:** Characteristics of Patients with Anxiety and Depression Diagnoses, (N=4805)

| **Patient Characteristics** | **n (%)** |
| --- | --- |
| Age group at first anxiety and/or depression visit, years |  |
| Under 2 | 35 (0.75) |
| 2-12 | 1873 (39.00) |
| 13-18 | 2826 (58.81) |
| 19-21 | 71 (14.78) |
| Mean age | 13.23 (SD = 3.82) |
| Male | 2005 (41.73) |
| Race |  |
| Asian | 488 (10.16) |
| Black or African American | 152 (3.16) |
| White or Caucasian | 1927 (40.10) |
| Other | 883 (18.38) |
| Unknown | 1355 (28.20) |
| Ethnicity |  |
| Hispanic | 543 (11.30) |
| Not Hispanic | 2712 (56.44) |
| Unknown | 1550 (32.26) |
| Insurance |  |
| Private | 3652 (76.00) |
| Public | 974 (20.27) |
| Military | 135 (2.80) |
| Unknown | 44 (0.92) |
| Diagnosis groups |  |
| Anxiety only (Anxiety) | 2635 (54.84) |
| Depression only (Depression) | 1433 (29.82) |
| Both anxiety and depression during study period (Anxiety+Depression) | 737 (15.34) |
| Comorbidities in patients with anxiety and depression |  |
| Attention-deficit/hyperactivity disorder | 876 (18.23) |
| Sleep disorders | 501 (10.43) |
| Trauma and stressor related disorders | 223 (4.64) |
| Autism spectrum disorder | 174 (3.62) |
| Obsessive compulsive and related disorders | 127 (2.64) |
| Feeding and eating disorders | 123 (2.56) |
| Disruptive, impulsive control, and conduct Disorders | 92 (1.91) |
| Substance-related and addictive disorders | 64 (1.33) |
| Bipolar and related disorders | 30 (0.62) |

SD = standard deviation

#### Comorbid Behavioral and Mental Health Conditions in Patients with Anxiety and/or Depression

Table 1 also includes the most common comorbid behavioral and mental health disorders: Attention-deficit/hyperactivity disorder (18.2%), sleep disorders (10.4%), trauma and stressor-related disorders (4.6%) and autism spectrum disorder (3.6%). 881 (33.4%) of patients with anxiety, 482 (33.6%) of patients with depression, and 365 (49.5%) of patients with anxiety+depression, had one or more co-existing behavioral or mental health condition.

#### Medications for Patients with Anxiety and/or Depression

Of 2635 patients with anxiety, 14% (n=370) received a psychoactive (non-stimulant) medication. Of 1433 children with depression, 20% (n=287) received a psychoactive medication. Of 737 children with anxiety+depression diagnoses during the study period, 47% (n=346) received a psychoactive medication.

Figure 1 summarizes most commonly used medications in patients with anxiety, depression, and anxiety+depression. The most common medications were all in the class of serotonin reuptake inhibitors (SSRIs). For patients with anxiety, the most commonly used medications were: 1) sertraline, 2) citalopram, and 3) fluoxetine. For patients with depression, the most commonly used medications were: 1) fluoxetine, 2) citalopram, and 3) sertraline. For patients with anxiety+depression, the most commonly used medications were: 1) citalopram, 2) sertraline, and 3) escitalopram.

**Figure 1:**
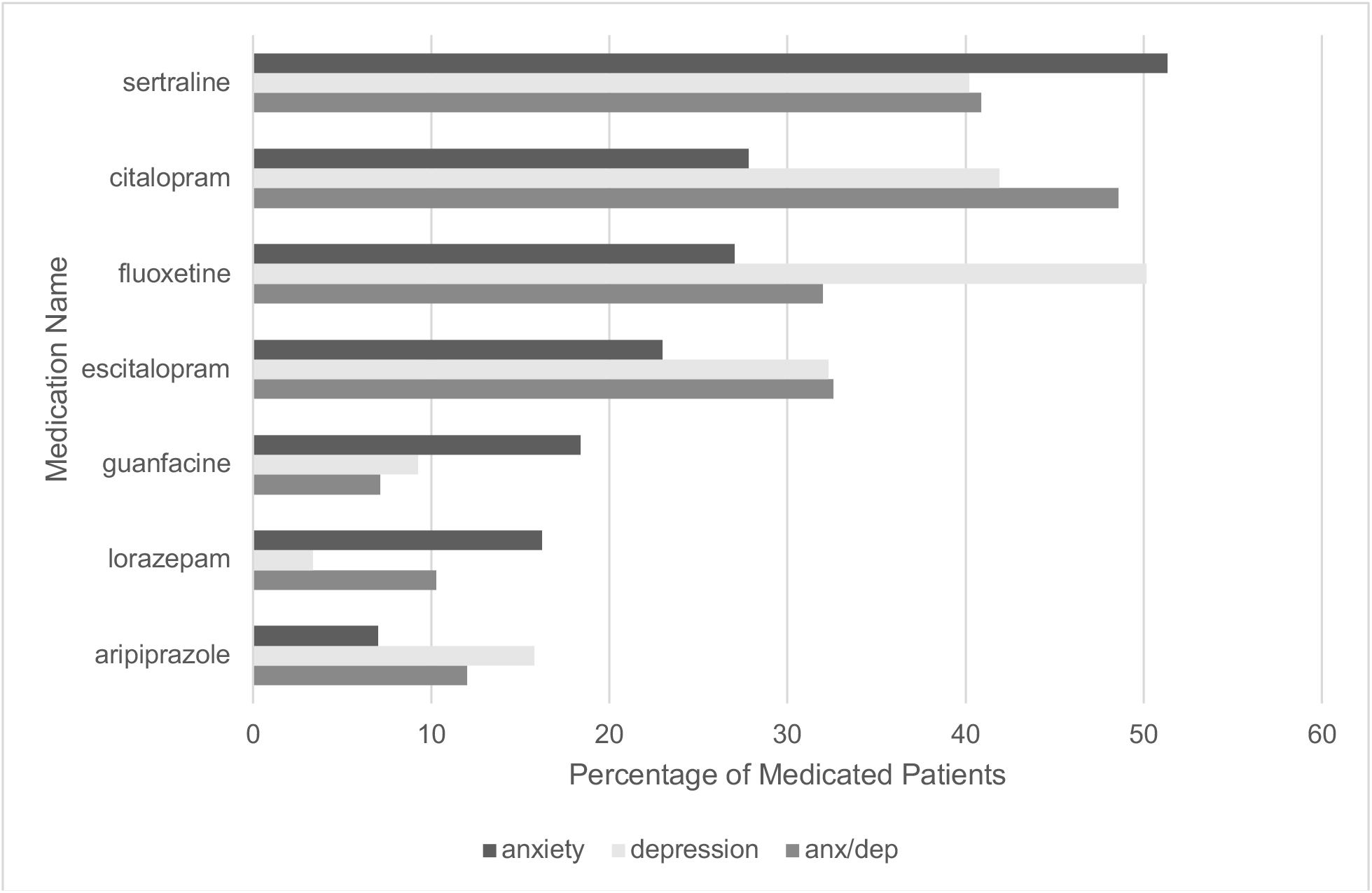
Most Common Medications for Anxiety, Depression, or Both.

Alprazolam, a benzodiazepine, appeared on the list of top ten most frequently prescribed medications for anxiety (7.0% of patients with anxiety) and lorazepam, another benzodiazepine, appeared on the top ten most frequently prescribed medications for anxiety+depression (10.3% of patients with anxiety+depression).

#### Medications for Patients with Symptom-Level Diagnoses

For patients with anxiety, none of the 51 patients with symptom-level diagnoses only, such as “worries”, received medication. For patients with depression, only 1 of the 216 patients (<1.0%) with symptom-level diagnoses only, such as “sadness”, received medication. For patients with anxiety+depression, 0 of the 2 patients with symptom-level anxiety and depression received medication.

#### Race

After implementing a chi-square analysis, we found that proportions of those medicated were not comparable across racial categories. A smaller proportion of Asian patients had medication prescription and a larger proportion of White had medication prescription (7.1% of Asian vs 15.8% of White for anxiety, 14% of Asian vs 22.2% of White for depression, and 28% of Asian vs 50.4% of White for anxiety+depression) (Supplement 4). There was no statistical difference in proportions of Hispanic and non-Hispanic patients who had medication prescription.

### Predictors of Medication Prescription, by Diagnosis Group: Crosstabs and Regression Models

Three logistic regression models were performed to determine patient factors associated with likelihood to be prescribed medication for patients with anxiety (Table 2), depression (Table 3), anxiety+depression (Table 4). Only patients 6-18 years were included in the models, as patients under 6 rarely received medication, and there were few patients over 18 in the dataset. See Supplement 5 for proportions of patients in each diagnostic category who received medication.

**Table 2:** Patient Characteristics Associated with Medication Prescription for Patients with Anxiety.

| Patient characteristics | Number of patients<br>(n)<br>(N=2385) | Medication Odds Ratio (95%<br>CI) | P value |
| --- | --- | --- | --- |
| Age |  |  |  |
| 6-12 | 1185 | Ref |  |
| **13-18** | **1200** | **2.26 (1.77-2.89)** | **<0.001** |
| Sex |  |  |  |
| Female | 1352 | 0.90 (0.70-1.15) | 0.41 |
| Insurance |  |  |  |
| Private | 1869 | Ref |  |
| Public | 425 | 1.11 (0.83-1.50) | 0.48 |
| Military | 65 | 0.57 (0.24-1.37) | 0.21 |
| NA | 26 | 0.54 (0.12-2.37) | 0.42 |
| Number of Comorbidities |  |  |  |
| 0 | 1570 | Ref |  |
| **1** | **637** | **2.47 (1.63-3.74)** | **<0.001** |
| **2+** | **178** | **4.70 (2.18-10.13)** | **<0.001** |
| Comorbid Diagnoses |  |  |  |
| ADHD | 4452 | 1.07 (0.69-1.66) | 0.78 |
| ASD | 104 | 1.64 (0.94-2.87) | 0.08 |
| **Sleep** | **227** | **0.50 (0.30-0.83)** | **0.007** |
| **Trauma and Stressor**<br>**Related Disorders** | **82** | **0.38 (0.18-0.80)** | **0.01** |
| Referral to DBP or Psychiatry | 232 | 1.15 (0.80-1.67) | 0.47 |

**\*\*** Nagelkerke R square: 0.10

**Table 3:**
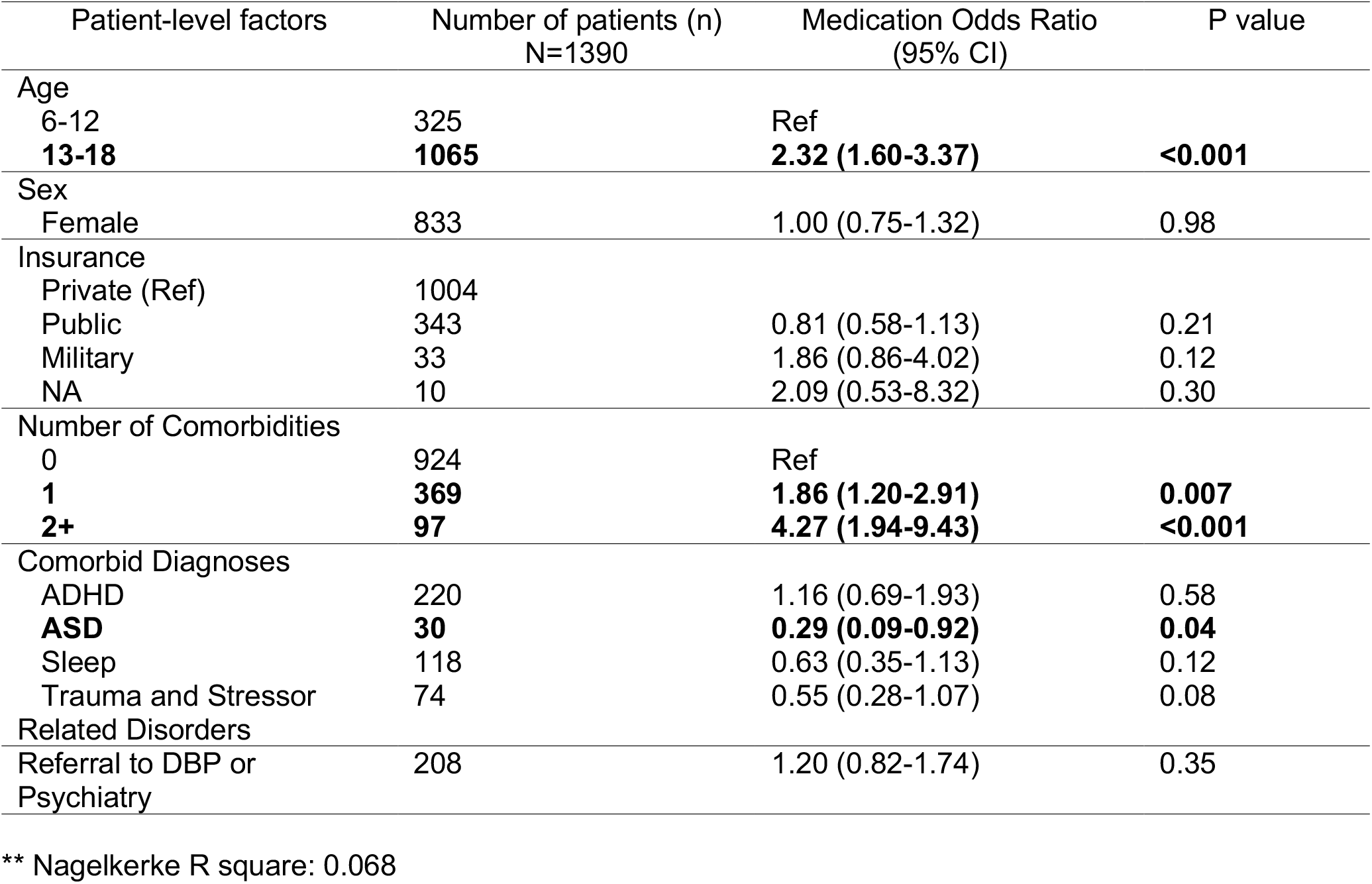
Patient Characteristics Associated with Medication Prescription for Patients with Depression.

**Table 4:**
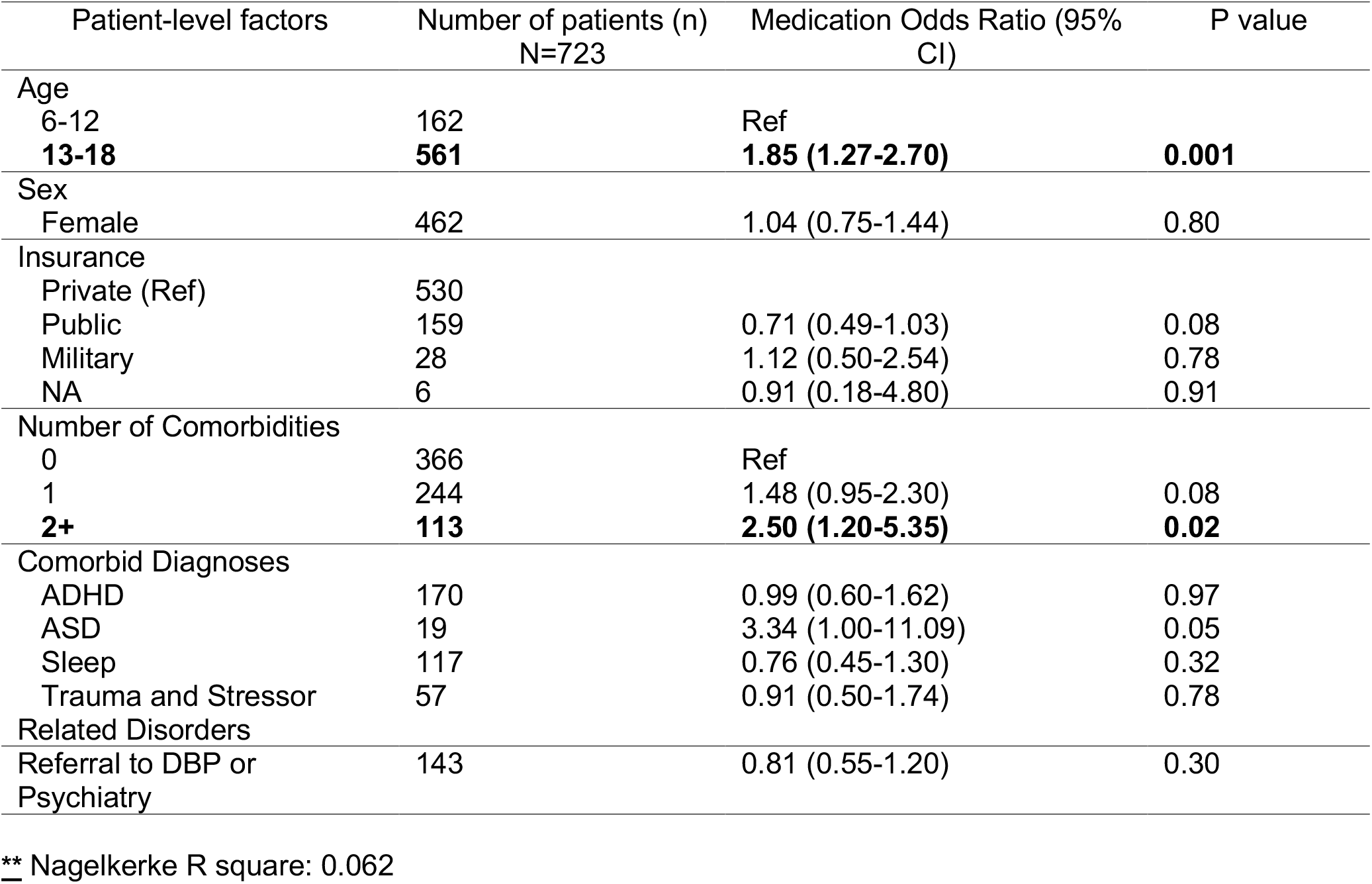
Patient Characteristics Associated with Medication Prescription for Patients with Anxiety and Depression.

#### Anxiety

For anxiety (Table 2), the model explained 10.0% (Nagelkerke *R*^*2*^) of the variance in medication prescription. For children with a diagnosis of anxiety, 13-18 year old children had 2.26 higher odds of receiving medication compared to 6-12 year old children (95% CI: 1.77-2.89, p<0.001). Children with behavioral and mental health comorbidities were more likely to receive medication than children without comorbidities ((one comorbidity: aOR 2.47, 95% CI: 1.63-3.74, p<0.001), (two or more comorbidities: aOR 4.70, 95% CI: 2.18-10.13, p<0.001)). Two specific comorbidities were associated with decreased likelihood of receiving medication: Comorbid Sleep Disorder (aOR: 0.50, 95% CI: 0.30-0.83, p=0.007), and Comorbid Trauma and Stressor Related Disorder (aOR: 0.38, 95% CI: 0.18-0.80, p=0.01). Comorbid ADHD and Autism Spectrum Disorder were not associated with medication prescription.

#### Depression

For depression (Table 3), the model explained 7.0% (Nagelkerke *R*^*2*^) of the variance in medication prescription. For children with a diagnosis of depression, 13-18 year old children had 2.32 higher odds of receiving medication compared to 6-12 year old children (95% CI: 1.60-3.37, p<0.001). Children with behavioral and mental health comorbidities were more likely to receive medication than children without comorbidities ((One comorbidity: aOR 1.86, 95% CI: 1.20-2.91, p=0.007), (Two or more comorbidities: aOR 4.27, 95% CI: 1.94-9.43, p<0.001)). Comorbid Autism Spectrum Disorder was associated with 0.29 lower odds of receiving medication compared to patients without an autism diagnosis (95% CI: 0.09-0.92, p=0.04). Comorbid ADHD, Sleep Disorders, and Trauma and Stressor Related Disorders were not associated with medication prescription.

#### Anxiety+Depression

For anxiety+depression (Table 4), model explained 5.5% (Nagelkerke *R*^*2*^) of the variance in medication prescription. For children with anxiety and depression, 13-18 year old children had 1.85 higher odds of receiving medication compared to 6-12 year old children (95% CI: 1.27-2.27, p=0.001). Children with two or more behavioral and mental health comorbidities had 2.50 higher odds of receiving medication compared with children with no comorbidities (95% CI: 1.20-5.35, p=0.02). Comorbid ADHD, Autism Spectrum Disorder, Sleep Disorders, and Trauma Stressor Related Disorders were not associated with medication prescription.

Across all models, patient sex, insurance type, and referral to DBP and/or psychiatry within Stanford Children’s Health were not associated with the likelihood of receiving a medication prescription.

## Discussion

We found that, within this population of children in Northern CA, 5.2% of patients had a diagnosis of anxiety and/or depression recorded in their primary care EHR. Comorbid developmental, behavioral, and mental health conditions were common in all three subcohorts. 33.4% of patients with anxiety, 33.6% of patients with depression, and 49.5% of patients with anxiety+depression had one or more co-existing behavioral or mental health conditions. Psychoactive medications were prescribed to 14% of patients with anxiety, 20.3% with depression, and 47.5% of patients with anxiety+depression.

For all three subcohorts (anxiety, depression, and anxiety+depression), patients with symptom-level (ICD-10 “R” codes) diagnostic codes rarely received medication. This finding suggests that providers do not prescribe medications in “mild” and/or diagnostically uncertain cases, which aligns with current guidelines for both anxiety and depression.^12–14^

Current literature indicates that SSRIs have the most evidence of beneficial effect in pharmacological treatment of anxiety and depression in pediatric patients. Clinical practice guidelines and practice parameters also support the use SSRI medications; fluoxetine and escitalopram recommended as first line for depression.^16^ High quality, NIH-sponsored multisite trials have supported safety and efficacy of fluoxetine.^16^ In our study, patients with anxiety or with depression most commonly were prescribed sertraline, citalopram, and fluoxetine. For patients with anxiety+depression, citalopram, sertraline, and escitalopram most commonly were prescribed. These findings indicate that the medications most commonly used by PCPs for anxiety and depression generally align with current evidence. However, a substantial proportion of patients who were prescribed medication during the study period received benzodiazepine medications at some point (32% for anxiety, 7% for depression, and 18% for anxiety+depression). Benzodiazepines have limited evidence for beneficial effect in pediatric patients.^15,18,21^ There are also concerns about safety and the potential of diversion.^22,23^ Further study is needed to understand the rationale for use of these medications by PCPs in this network. In particular, it would be important to determine if subspecialist involvement was associated with prescription of benzodiazepines in these pediatric patients.

We investigated patient factors associated with likelihood to be prescribed medication with separate models for 1) anxiety, 2) depression, and 3) anxiety+depression. The models accounted for 5-10% of the variance in medication use. Thus, it appears that other factors not assessed here are important to the decision to treat with medication. For all three subcohorts, older age and higher number of comorbidities were associated with increased likelihood to receive medication. For patients with anxiety, comorbid sleep disorder or trauma and stressor related disorder were independently associated with <u>decreased</u> likelihood to be prescribed medication, suggesting that clinicians may have a different clinical approach -- perhaps focusing more on non-pharmacological strategies -- for patients with those co-morbidities.

This study has several limitations. First, race and ethnicity data are missing from a significant percentage of the patient records, as these data are not routinely collected in certain practices within the network. Of the complete data, we found that, compared to patients of other race groups, White patients were more likely and Asian patients were less likely to be prescribed medication across all diagnostic groups (7.1% of Asian vs 15.8% of White for anxiety, 14% of Asian vs 22.2% of White for depression, and 28% of Asian vs 50.4% of White for anxiety+depression). A growing body of literature indicates that Asian American patients experience significant barriers (found in healthcare systems and/or family systems) to accessing mental healthcare.^24–26^ Thus, further investigation of this finding is warranted.

Next, we were not able to consider the effects of referral to external psychiatry resources or referral to psychotherapy as this information was not captured in the electronic health record structured data. A more thorough understanding of PCP recommendation of non-pharmacologic treatments to these children will require chart review. Finally, although the primary care network from which our data was obtained is community-based, the patient demographics are not representative of demographics in California (our data set includes higher percentage white and private insurance), limiting the generalizability of the findings.^27^

## Conclusions

This study of a large community-based network of pediatrics practices showed that medications prescribed by primary care providers for anxiety and depression generally align with current recommendations. The most commonly used medications were SSRIs. Further study is needed to understand why providers use benzodiazepines, which have less evidence for beneficial effect, and more concern for side effects, diversion, and unintended recreational use. PCPs prescribe medications more frequently for patients with anxiety+depression than for patients with either diagnosis alone. Older age and higher number of comorbidities were associated with increased likelihood to be prescribed medication. Further study also is needed to investigate how PCPs collaborate with subspecialists in the care of patients with anxiety and depression.

## Data Availability

The authors confirm that the data supporting the findings of this study are available within the article [and/or] its supplementary materials.

