## Supplemental Figures for "Medication Management of Anxiety and Depression by Primary Care Pediatrics Providers: A Retrospective Electronic Health Record Study"

### Supplement 1: Cohort Selection Diagram

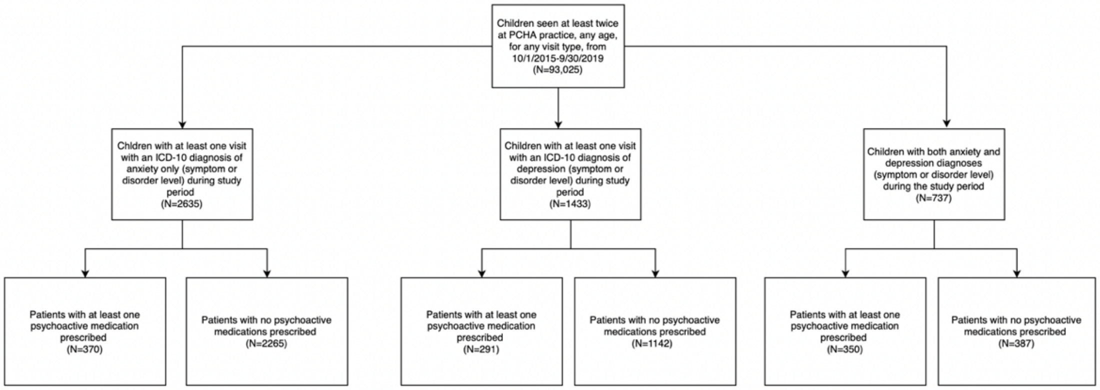

### Supplement 2 : ICD 10 Codes and Text descriptors Used for Study Outcomes

| Condition | ICD-10 Diagnosis Code | ICD-10 Text Descriptors |
|---|---|---|
| Anxiety | | |
| Symptom Level | R45.2 | Unhappiness (worries NOS) |
| | R45.82 | Worries |
| | R45.89 | Feeling worried, anxious appearance, feeling anxious |
| Disorder Level | F06.4,F10.180,F10.280,F12.980,F13.180,F13.280,F13.980,F14.180,F14.280,F14.980,F15.180,F15.280,F15.980,F16.180,F16.280,F16.980,F18.180,F18.280,F18.980,F19.180,F19.280,F19.980,F31.3,F40.0,F40.00,F40.01,F40.02,F40.10,F40.11,F40.210,F40.218,F40.220,F40.228,F40.23,F40.230,F40.231,F40.233,F40.240,F40.241,F40.242,F40.243,F40.248,F40.291,F40.298,F40.8,F40.9,F41.0,F41.1,F41.3,F41.8,F41.9,F93.0,F94.0 | All |
| Depression | | |
| Symptom Level | R45.7 | State of emotional stress or shock |
| | R45.81 | Low self esteem |
| | R45.84 | Anhedonia |
| | R45.851 | Suicidal ideations |
| | R45.86 | Emotional lability |
| | R45.89 | Sad, sad mood, feeling sad, sadness, feeling of sadness, at risk for self harm, depressed affect, difficulty coping, dysphoric mood, feelings of worthlessness, feels depressed, flat affect, non-suicidal depressed mood, non-suicidal self harm, self-esteem disturbance, suicidal risk, tearfulness, thoughts of self harm |
| Disorder Level | F06.30,F06.31,F06.32,F06.34,F10.14,F10.24,F10.94,F11.14,F11.24,F11.94,F13.14,F13.24,F13.94,F14.14,F14.24,F14.94,F15.14,F15.24,F15.94,F16.14,F16.24,F16.94,F18.14,F18.24,F18.94,F19.14,F19.24,F19.94,F32.0,F32.1,F32.2,F32.3,F32.4,F32.5,F32.8,F32.81,F32.89,F32.9,F33.0,F33.1,F33.2,F33.3,F33.40,F33.41,F33.42,F33.8,F33.9,F34.1,F34.81,F34.89,F34.9 | All |

### Supplement 3: Medication Prevalence By Age

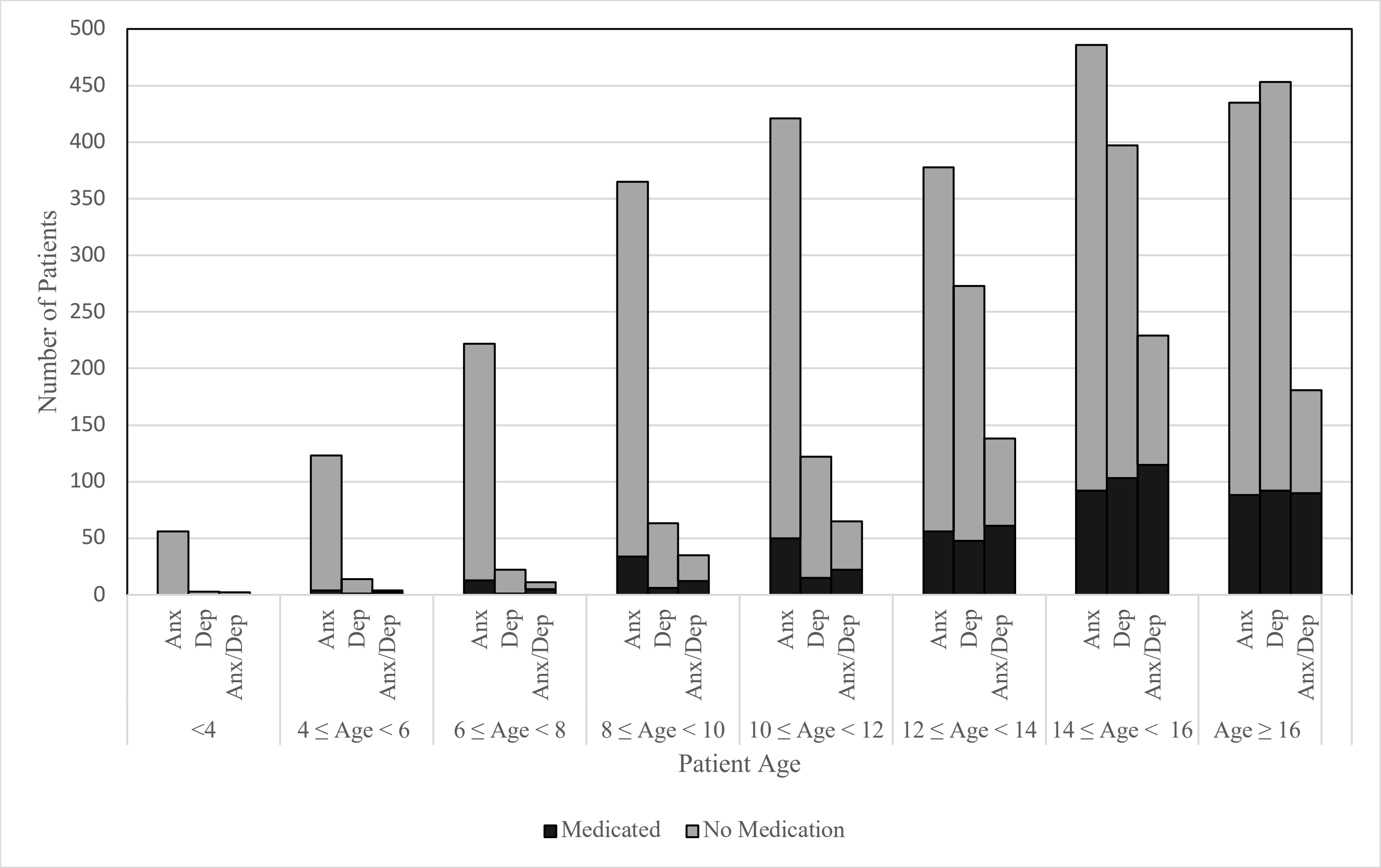

### Supplement 4: Medication Prevalence by Race and Ethnicity

| Diagnostic Category | Ethnicity | | Race | | | |
|---|---|---|---|---|---|---|
| | Hispanic | Non-Hispanic | Asian | Black | White | Other |
| Anxiety (# Medicated/Total) (%) | 30/265 (11.3%) | 178/1409 (12.6%) | 17/241 (7.1%) | 7/72 (9.7%) | 160/1011 (15.8%) | 37/427 (8.7%) |
| Depression (# Medicated/Total) (%) | 24/175 (13.7%) | 141/756 (18.7%) | 22/157 (14%) | 6/45 (13.3%) | 117/527 (22.2%) | 34/268 (12.7%) |
| Anxiety+Depression (# Medicated/Total) (%) | 34/83 (41.0%) | 155/356 (43.5%) | 14/50 (28%) | 11/23 (47.%) | 134/266 (50.4%) | 52/139 (37.4%) |

### Supplemental Figure 5: Medication Prevalence by Patient Factor for 6-18 year old patients

| | Anxiety (N=2385) | | Depression (N=1390) | | Anxiety+Depression (N=723) | |
|---|---|---|---|---|---|---|
| | Medicated | Nonmedicated | Medicated | Nonmedicated | Medicated | Nonmedicated |
| Age | | | | | | |
| 6-12 | 125/1185 (10.54%) | 1060/1185 (89.45%) | 40/325 (12.30%) | 285/325 (87.70%) | 61/162 (37.65%) | 101/162 (62.35%) |
| 3-18 | 232/1200 (19.33%) | 968/1200 (80.67%) | 245/1065 (23.00%) | 820/1065 (77.00%) | 282/561 (50.26%) | 279/561 (49.74%) |
| Sex | | | | | | |
| Female | 184/1352 (13.60%) | 1168/1352 (86.40%) | 166/833 (19.92%) | 667/833 (80.08%) | 218/462 (47.18%) | 244/462 (52.82%) |
| Male | 173/1033 (16.74%) | 860/1033 (83.26%) | 119/557 (21.36%) | 438/557 (78.64%) | 125/261 (47.89%) | 136/261 (52.11%) |
| Insurance | | | | | | |
| Private | 280/1869 (14.98%) | 1589/1869 (85.02%) | 212/1004 (21.12%) | 792/1004 (78.88%) | 263/530 (49.62%) | 267/530 (50.38%) |
| Public | 69/425 (16.23%) | 356/425 (83.77%) | 59/343 (17.21%) | 284/343 (82.79%) | 64/159 (40.25%) | 95/159 (59.75%) |
| Military | 6/65 (9.23%) | 59/65 (90.77%) | 11/33 (33.33%) | 22/33 (66.67%) | 13/28 (46.42%) | 15/28 (53.57%) |
| NA | 2/26 (7.70%) | 24/26 (92.30%) | 3/10 (30.00%) | 7/10 (70.00%) | 3/6 (50.00%) | 3/6 (50.00%) |
| Number of Co-morbidities | | | | | | |
| 0 | 170/1570 (10.83%) | 1400/1570 (89.17%) | 160/924 (17.32%) | 764/924 (82.68%) | 152/366 (41.53%) | 214/366 (58.47%) |
| 1 | 133/637 (20.88%) | 504/637 (79.12%) | 92/369 (24.94%) | 277/369 (75.06%) | 123/244 (50.40%) | 121/244 (49.60%) |
| 2+ | 54/178 (30.34%) | 124/178 (69.66%) | 33/97 (34.03%) | 64/97 (65.97%) | 68/113 (60.18%) | 45/113 (39.82%) |
| Comorbid Diagnoses | | | | | | |
| ADHD | 116/452 (25.66%) | 336/452 (74.34%) | 69/220 (31.37%) | 151/220 (68.63%) | 93/170 (54.71%) | 77/170 (45.29%) |
| ASD | 40/104 (38.46%) | 64/104 (61.54%) | 4/30 (13.33%) | 26/30 (86.67%) | 15/19 (78.94%) | 4/19 (21.06%) |
| Sleep | 38/227 (16.74%) | 189/227 (83.26%) | 27/118 (22.88%) | 91/118 (77.12%) | 60/117 (51.28%) | 57/117 (48.72%) |
| Trauma and Stressor Related Disorders | 10/82 (12.20%) | 72/82 (87.8%) | 16/74 (21.62%) | 58/74 (78.38%) | 30/57 (52.63%) | 27/57 (47.37%) |
| Referral to DPB or Psychiatry | 43/232 (18.54%) | 189/232 (81.46%) | 48/208 (23.08%) | 160/208 (76.92%) | 63/143 (44.05%) | 80/143 (55.95%) |
